# Estimating event ban effects on COVID-19 outbreak in Japan

**DOI:** 10.1101/2020.12.29.20248977

**Authors:** Junko Kurita, Tamie Sugawara, Yasushi Ohkusa

## Abstract

**Background:** Before Olympic and Paralympic Games in Tokyo, whether an audience shall be allowed or not has been a subject of concern in Japan as of early June, 2021. Object: We evaluated effects of professional baseball games with audiences as an example of the large sports events, on COVID-19 infectiousness.

**Method:** We regressed the effective reproduction number R(*t*) on a dummy variable for professional baseball games with audiences as along with temperature, humidity, mobility, and countermeasures. We examined two study periods: those including and excluding before initiation of the games in 2020.

**Results:** Estimation results indicate that the period with audiences exhibited significantly lower infectiousness than when audiences were excluded before initiation of the games with audience attendance. However, audiences were found to have a negative but insignificant effect when compared to the period before initiation of the attended games.

**Discussion and Conclusion:** This study found no clear evidence indicating that big sports events with audiences raise the COVID-19 infectiousness.

## Introduction

The Olympic and Paralympic Games in Tokyo have been planned to commence on July 23, 2021. Whether audiences will be allowed to attend game events or not has been the subject of great concern in Japan as of the beginning of June, 2021 [1].

As countermeasures against the COVID-19 outbreak in Japan, school closure and voluntary event cancellation were enacted from February 27, 2020 through the end of March. Large commercial events were cancelled. Subsequently, a state of emergency was declared for April 7 through 25 May, stipulating voluntary restrictions against leaving home. Consumer businesses such as retail shops and restaurants were shut down. During this period, the first peak of infection was reached on April 3. Infections subsequently decreased until July 29. The so-called “Go To Travel Campaign” (GTTC) started on July 22 as a 50% subsidized travel program aimed at supporting sightseeing businesses with government-issued coupons for use at shopping at tourist destinations. It was expected that the campaign might expand the outbreak. Thereafter, GTTC continued to the end of December, by which time a third wave of infection had emerged. The third wave in December, which was larger than either of the preceding two waves, reached its highest peak at the end of December. Therefore, GTTC was inferred as the main reason underlying the third wave [1].

To force the third wave lower, the second emergency status was declared on January 8, 2021 to March 15, 2021. However, the fourth wave emerged probably because of the spread of variant strains at the end of February. Moreover, to support the hosting of the Olympics and Paralympics games in Tokyo in July, a third emergency state was declared on April 25, 2021.

Nevertheless, although results were mixed, some results of studies suggest that COVID-19 might be associated with climate conditions, at least in China [2–4]. If that were true for Japan, then GTTC might not be the main reason for the third wave.

Moreover, mobility was inferred as the main cause of the outbreak dynamics, at least in the first wave in Japan [5] and throughout the world [6]. Therefore, for this study, we evaluate the GTTC effects on infectiousness and the effective reproduction number R(t) while considering climate conditions and mobility.

The object of this study was evaluation of the effects of professional baseball games with audiences as examples of big sports events, to infectiousness in COVID-19.

## Methods

The numbers of symptomatic patients reported by the Ministry of Health, Labour and Welfare (MHLW) for January 14 – May 4, published [7] as of May 21, 2021 were used. Some patients were excluded from data for Japan: those presumed to be persons infected abroad or infected as Diamond Princess passengers. Those patients were presumed not to represent community-acquired infection in Japan. For onset dates of some symptomatic patients that were unknown, we estimated their onset date from an empirical distribution with duration extending from the onset to the report date among patients for whom the onset date had been reported.

The following procedure is similar to that used for our earlier research [8,9]. As described hereinafter, we estimated the onset date of patients for whom onset dates were not reported. Letting *f*(*k*) represent this empirical distribution of incubation period and letting *N*_*t*_ denote the number of patients for whom onset dates were not available published at date *t*, then the number of patients for whom the onset date was known is *t*-1. The number of patients with onset date *t*-1 for whom onset dates were not available was estimated as *f*(1)*N*_*t*_. Similarly, patients with onset date *t*-2 and for whom onset dates were not available were estimated as *f*(2)*N*_*t*_. Therefore, the total number of patients for whom the onset date was not available, given an onset date of *s*, was estimated as Σ_*k*=1_*f*(*k*)*N*_*s*_+*k* for the long duration extending from *s*.

Moreover, the reporting delay for published data from MHLW might be considerable. In other words, if *s*+*k* is larger than that in the current period *t*, then *s*+*k* represents the future for period *t*. For that reason, *N*_*s*+*k*_ is not observable. Such a reporting delay engenders underestimation of the number of patients. For that reason, it must be adjusted as Σ_*k=1*_^*t-s*^*f*(*k*)*N*_*s*_+*k*/Σ_*k=1*_^*t-s*^*f*(*k*). Similarly, patients for whom the onset dates were available are expected to be affected by the reporting delay. Therefore, we have *M*_*s*_|_*t*_/Σ_*k=1*_^*t-s*^*f*(*k*), where *M*_*s*_|_*t*_ represents the reported number of patients for whom onset dates were period *s* as of the current period *t*.

We defined R(*t*) as the number of infected patients on day *t* divided by the number of patients who were presumed to be infectious. The number of infected patients was calculated from the epidemic curve by the onset date using an empirical distribution of the incubation period, which is Σ_*k*=*1*_*f*(*k*)*E*_*t+k*_, where *E*_*t*_ denotes the number of patients for whom the onset date was period *t*. The distribution of infectiousness in symptomatic and asymptomatic cases *g*(*k*), was assumed to be 30% on the onset day, 20% on the following day, and 10% for the subsequent five days [10]. Then the number of infectiousness patients was Σ_*k*=*1*_*g*(*k*)*E*_*t*-*k*_. Therefore, R(*t*) was defined as Σ_*k*=*1*_*f*(*k*)*E*_*t+k*_/Σ_*k*=*1*_*g*(*k*)*E*_*t*-*k*_.

We use average temperature and relative humidity data for Tokyo during the day as climate data because national average data were not available. We obtained data from the Japan Meteorological Agency (https://www.data.jma.go.jp/gmd/risk/obsdl/index.php). We have identified several remarkable countermeasures in Japan: two emergency status declarations, GTTC, and school closure and voluntary event cancellation (SCVEC). The latter, SCVEC, extended from February 27 through March: this countermeasure required school closure and cancellation of voluntary events, including private meetings. Then the first state of emergency was declared April 7. It ceased at the end of May. It required voluntary restriction against going out, school closures, and shutdown of businesses. To subsidize travel and shopping at tourist destinations, GTTC started on July 22 and ceased temporarily at the end of December. The second state of emergency was declared on January 7, 2021 for the 11 most affected prefectures. This countermeasure required restaurant closure at 8:00 p.m. and voluntary restriction against going out, but did not require school closure. It will continue until March 21, 2021. The third state of emergency was declared on April 25, 2021 for 4 prefectures, Tokyo, Osaka, Hyogo, and Kyoto prefectures. Then, application areas were extended gradually, but did not cover the entirety of Japan.

To clarify associations among R(*t*) and climate, mobility, and countermeasures, we regressed the daily R(*t*) on dummy variable for games with audience, daily climate, mobility, and countermeasures using ordinary least squares. A dummy variable for games with audience was defined as one after July 10, 2020 when limited audience was allowed until 5,000 audiences. Temperatures were measured in degrees Celsius, humidity, and mobility as percentages in regression, not as standardized. When some variables were found to be not significant, we excluded them and estimated the regression line again. If some variables were not significant in the full specification estimation, then we estimated it again without those non-significant variables, stepwisely.

We examine two study periods: the longer one including the duration before the season started, extending from February 1 through November 25, 2020, when the post season match of the professional baseball games in Japan was finalized. The shorter one was from June 19, when the season started, to November 25. We adopted 5% as the significance level.

## Results

Figure 1 presents an empirical distribution of the duration of onset to reporting in Japan. The maximum delay was 31 days. Figure 2 depicts an empirical distribution of incubation periods among 91 cases for which the exposed date and onset date were published by MHLW in Japan. The mode was six days. The average was 6.6 days.

**Figure 1:**
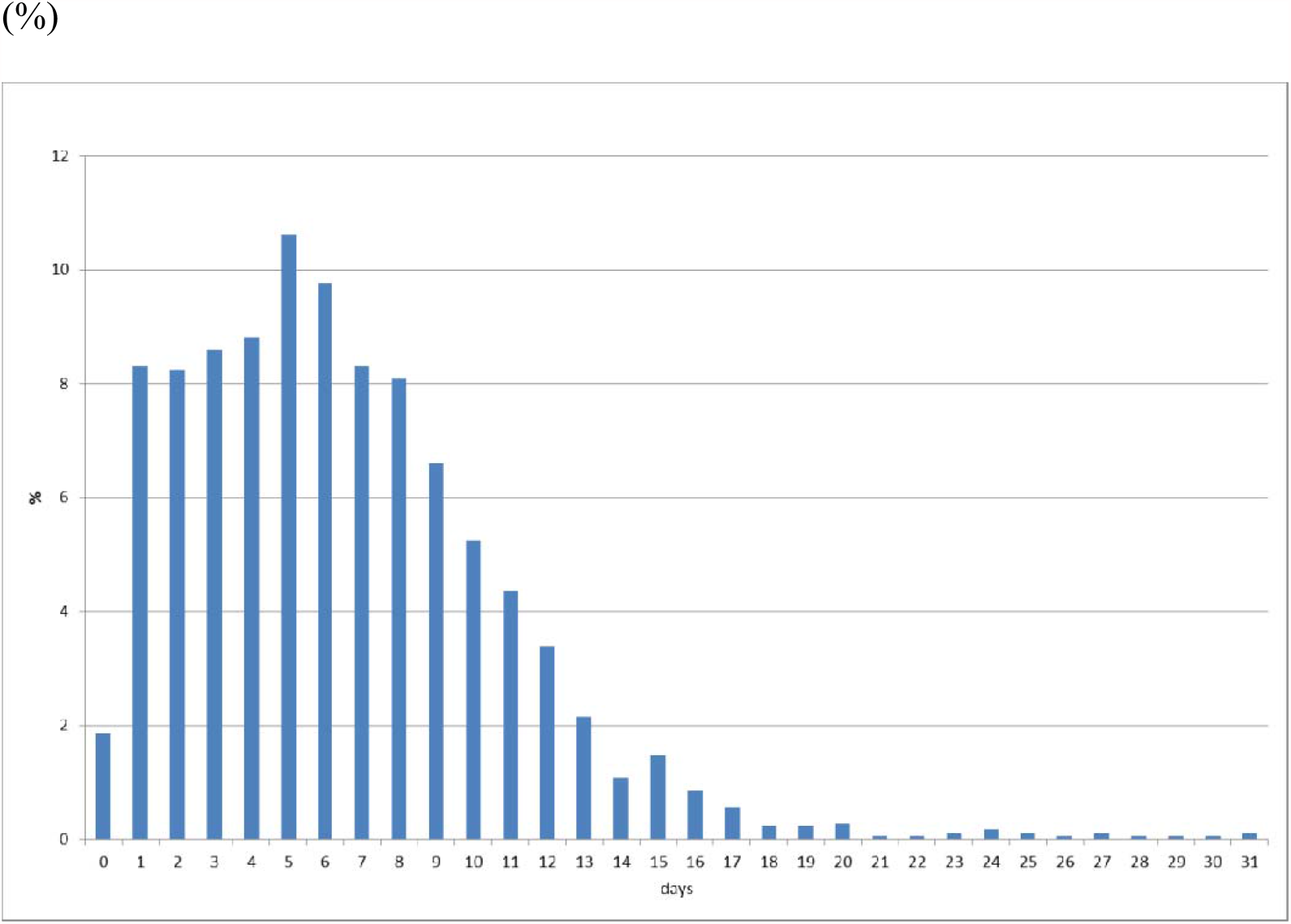
Empirical distribution of duration from onset to report by MHLW, Japan. Note: Bars represent the probability of duration from onset to report based on 657 patients for whom the onset date was available in Japan. Data were obtained from MHLW, Japan.

**Figure 2:**
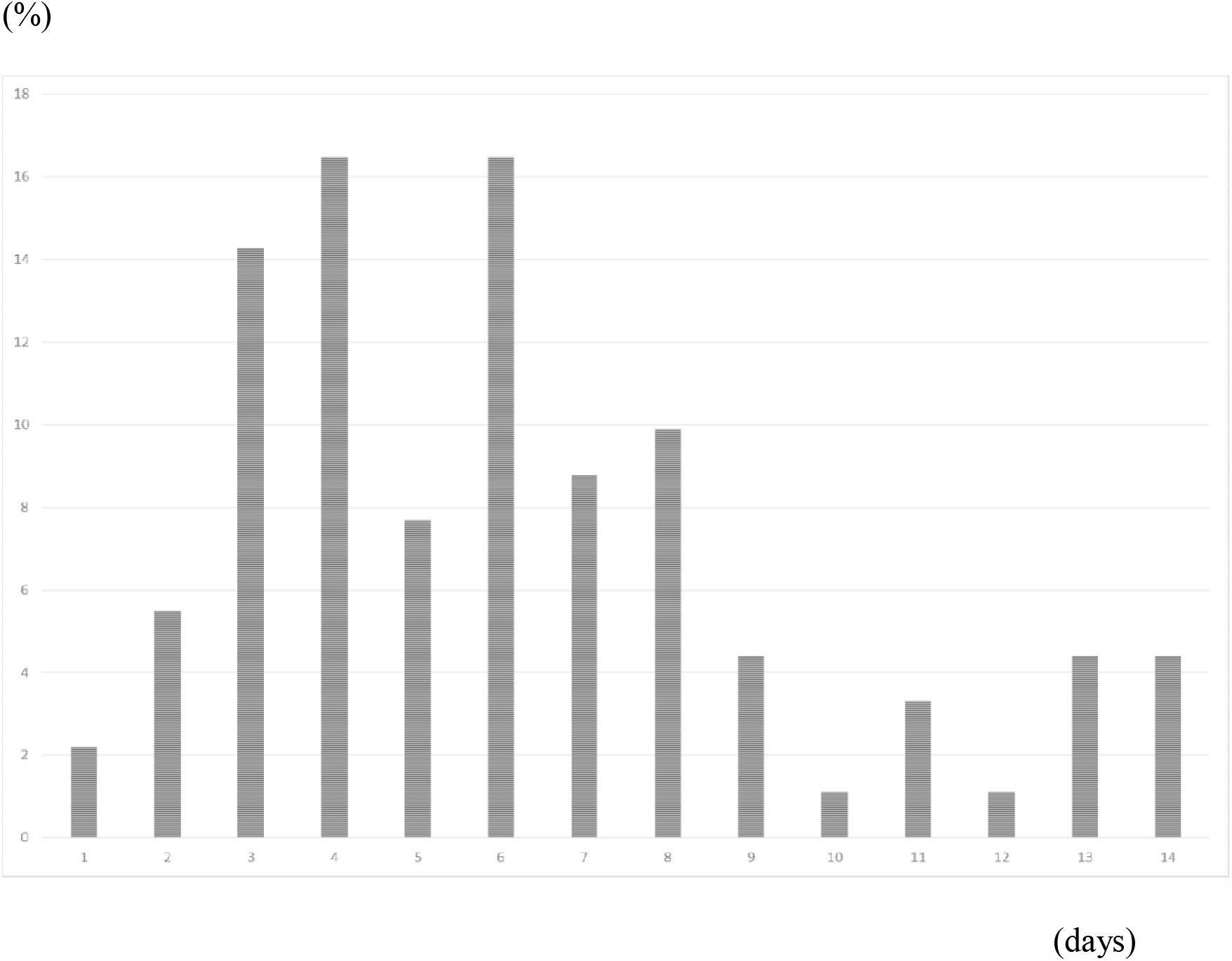
Empirical distribution of the incubation period published by MHLW, Japan. Notes: Bars show the distribution of incubation periods for 91 cases for which the exposure date and onset date were published by MHLW, Japan. Patients for whom incubation was longer than 14 days are included in the bar shown for day 14.

Table 1 presents estimation results including those obtained before the season. The left-hand side of Table 1 presents estimation results for the full specification with climate condition as an explanatory variable. The four columns at the middle of the table show estimation results without temperature or humidity because these were not significant in the full specification on the left-hand side. Neither temperature nor humidity was found to be significant. Therefore, we also estimated results without climate conditions and show the result on the right-hand side of the table.

**Table 1:**
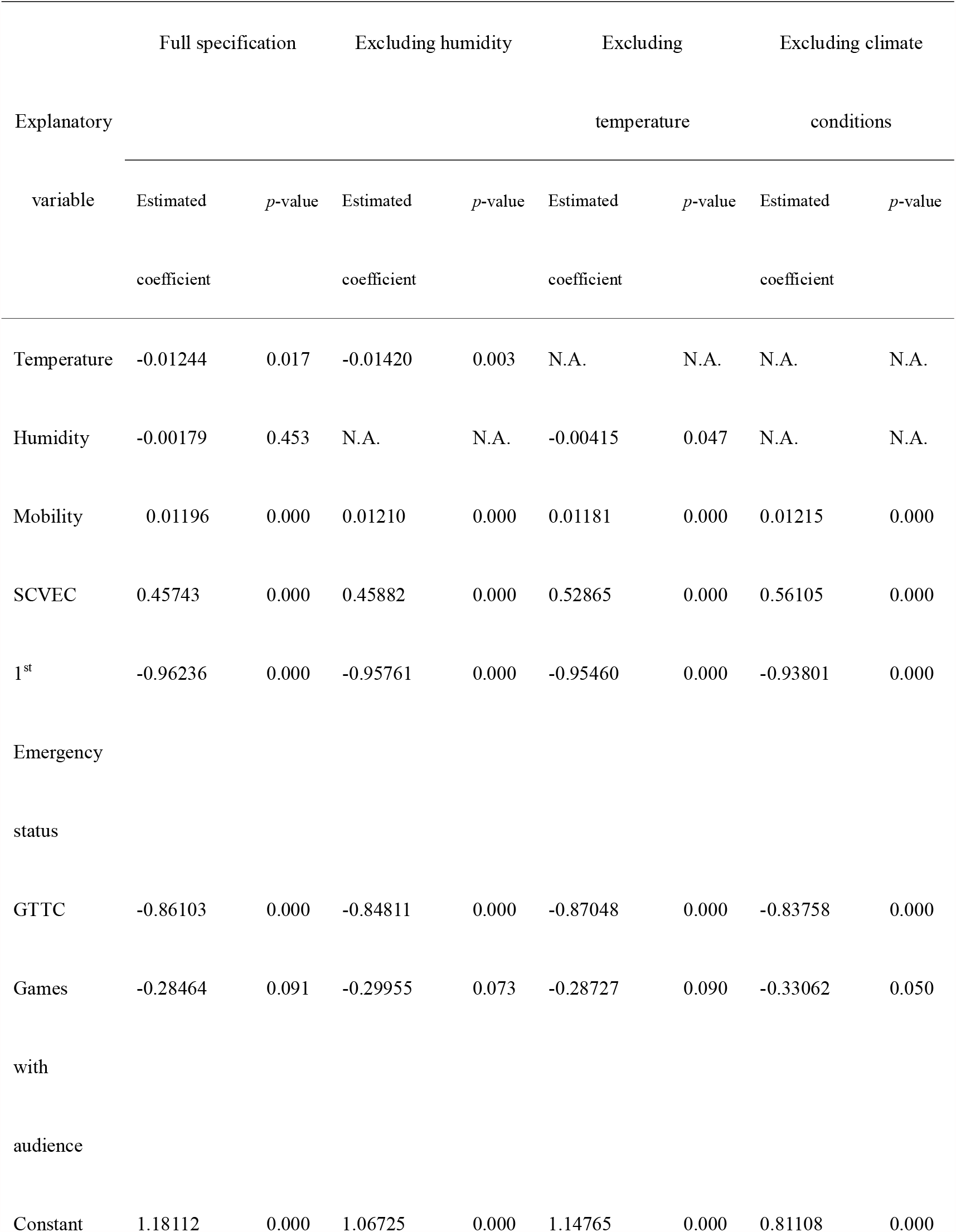
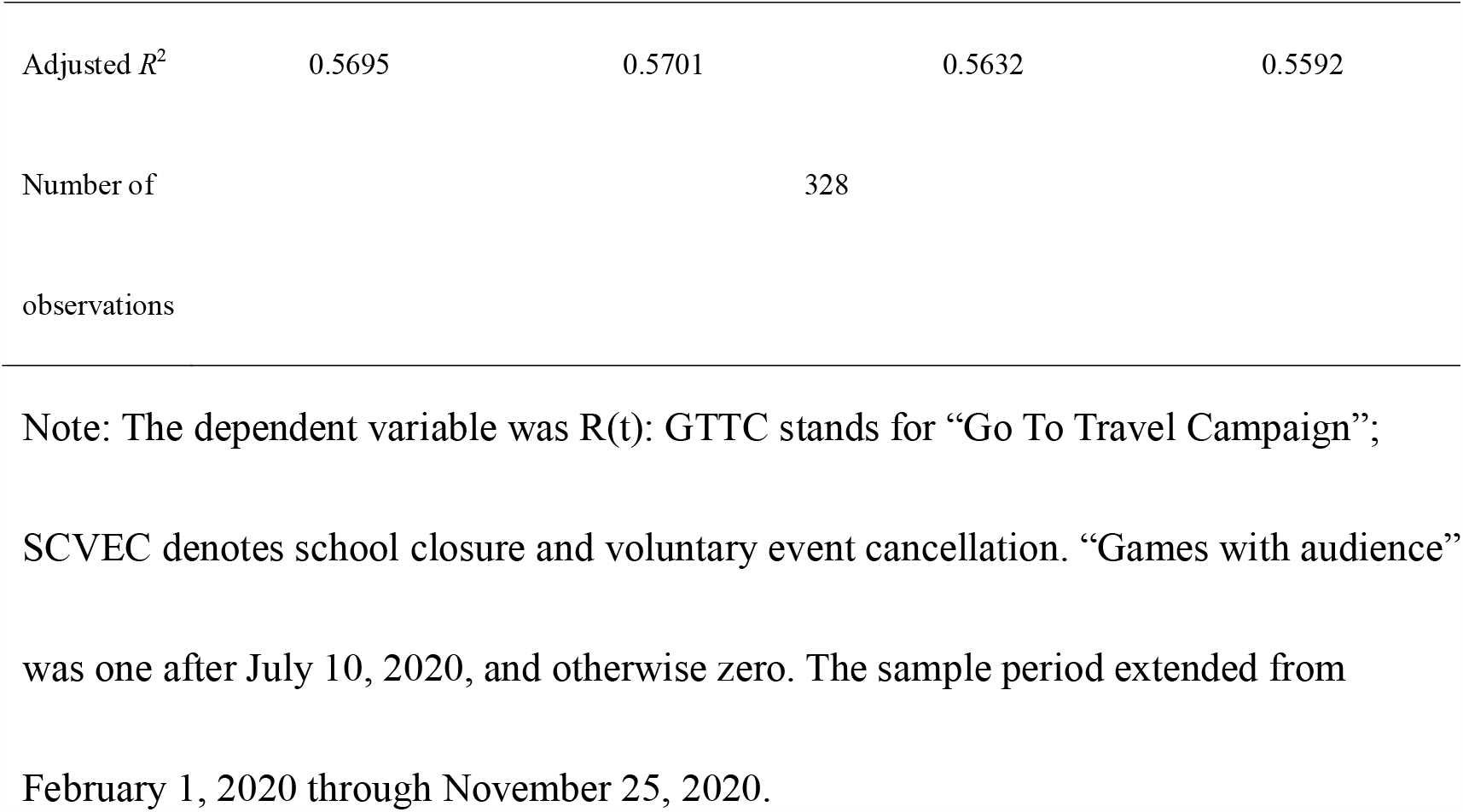
Estimation results of estimate R(t) for the climate condition, mobility, and countermeasures under full specifications, and excluding temperature and humidity

Mobility was positive and significant. Humidity was not significant when temperature was included. The first emergency status and GTTC were found to be negative and significant, but SCVEC was significant and positive. Games with an audience were negative but not significant for all specifications, even though it was marginally significant when climate conditions were excluded.

Table 2 presents estimation results in the shorter study period. In this case, although humidity was similar with Table 1, mobility was not significant when climate conditions were excluded. Games with an audience were significant and negative in all specifications in Table 2.

**Table 2:**
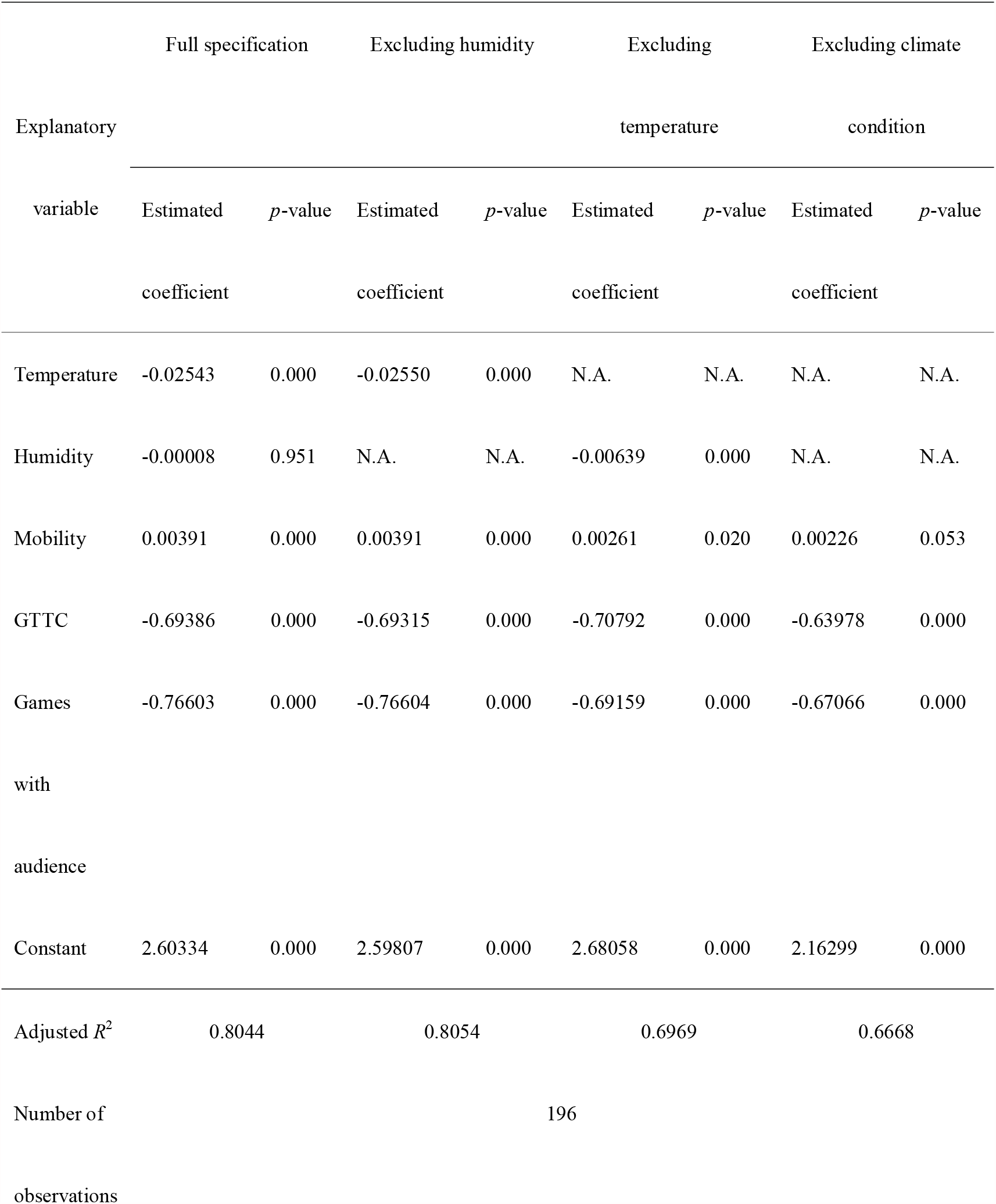
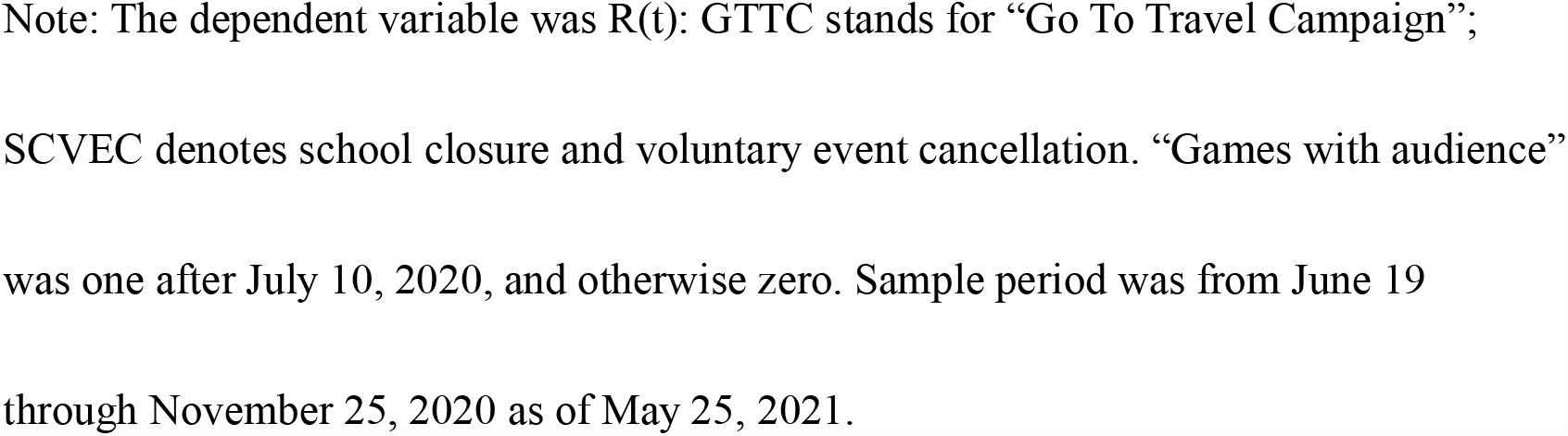
Estimation results of estimate R(t) for climate conditions, mobility, and countermeasures under the full specification and excluding temperature and/or humidity during period of Professional Baseball Games in 2020: June 19 to November 25, 2020

## Discussion

Estimation results showed that games with an audience was significantly negative in a shorter study period but not in longer period. At least, one can infer that it did not raise infectiousness. Results suggest that GTTC also reduced R(*t*) by about the same magnitude. Many observers have presumed that GTTC raised infectiousness, but it might have instead reduced infectiousness.

This counterintuitive result about GTTC might reflect the situation for July. Before GTTC was started on July 22, even though temperatures were high, a second peak occurred, indicating that the non-GTTC period had been affected by high infectiousness. By contrast, in November or December, R(*t*) was not so high as either the first and second wave. Of course, temperatures then were lower than they had been for either of the prior two peaks.

It is noteworthy that R(*t*) does not represent the number of newly infected patients. It is a ratio divided by the number of patients with infectiousness. The number of newly infected patients in the third wave around the end of 2020 was much higher than either of the two prior waves, as shown in Figure 1. Therefore, the greater number of patients in the third wave resulted from the greater number of patients with infectiousness, and not larger R(*t*). Larger numbers of patients with infectiousness were reported because of the fact that R(*t*) was not so high, but higher than any for a long time: about three months. One must be reminded for interpretation of the obtained result that climate conditions, mobility, and countermeasures affect infectiousness R(*t*), but they do not directly affect the number of patients.

Reportedly [11], travel-associated COVID-19 incidence during July 22–26, when GTTC started, was much higher than during either the earlier period of June 22 to July 21 or July 15–19 or June 22 – July 21 in terms of the incidence rate ratio (IRR). That earlier study also compares the period of August 8–31. Patient data of two types were used: the onset date and the date of a positive test result.

We have identified some odd points in the report of that study. The first is that the proportion of people with a travel history during the GTTC period was comparable to those of people during the two prior periods. Especially, when the earlier period was defined as July 15–19, the proportion of people with a travel history among patients with an available onset date was smaller for the GTTC period than during the prior period. However, the authors of that report found significantly higher incidence at the time when GTTC started. The findings of that other study might merely reflect the fact that the total number of patients in the GTTC period was higher than during the prior period. In other words, they did not control the underlying outbreak situation and therefore found incorrect association. Use of the IRR would be valid if the underlying outbreak situation other than the examining point was the same in the two considered periods. Therefore, application of IRR might be inappropriate for this issue. At least, controlling the potential differences in the outbreak situation is expected to be necessary.

The underlying outbreak situation, unrelated to GTTC, was reflected in the number of patients without a travel history or any sightseeing. To control the underlying outbreak situation, analysis of the share of patients with a travel history or sightseeing might be one procedure. However, that share did not increase markedly during the GTTC starting period. This fact indicates that the results and conclusions from that earlier study are misleading.

A second point is that the authors of that report referred to the period of August 8–31, when GTTC was continuing. The proportion of patients with a travel history or tourism was much smaller than in the GTTC period or in the prior period. Although the authors did not compare incidence in the period with that of either the prior period or the GTTC period, the rate of incidence during the period in August was probably lower than in other periods. In fact, some patients practicing GTTC might have been included in the period, as described above. Their inclusion might be inconsistent with the authors’ conclusion.

A third point is that, as shown in Figure 1, which was created using the same procedure as that reported for earlier studies [8,9], we used publicly reported information [7] to ascertain the peak of newly infected persons as July 23: the GTTC starting date. Therefore, we infer that GTTC possibly reduced infectiousness. We also consider climate conditions. At around the end of July, the rainy season in Japan ceased; summer began, bringing with it high temperatures. At least, GTTC was insufficient to raise the number of patients and cancel out benefits from the improved climate conditions. Taken together, these points suggest that GTTC might not be the main factor affecting the course of the outbreak.

Moreover, GTTC must increase the number of patients without a travel history if GTTC has a strong effect on the outbreak. For example, a patient travelling while using GTTC on July 22 and 23 and then showing onset on July 24 had a travel history with GTTC, but would not be included in a group of patients with a travel history whose onset date was included in the GTTC start period of July 27–31. However, presymptomatic patients are known to be infectious during the symptomatic period [12]. Such a patient might infect hotel staff members or persons in visiting areas. They did not have a travel history. Their onset dates were July 27 and 28. Actually, they included a group of patients with no travel history in the GTTC starting period of July 27–31. Therefore, GTTC certainly increased the number of patients without a travel history, but it did not increase patients with a travel history in this case. For that reason, when considering GTTC effects, the number of patients must be checked irrespective of their travel history.

The third wave of the outbreak, much larger than the second wave which struck around July, showed its peak as around the end of the year. Almost simultaneously, GTTC ceased on December 28. These two facts imply that stopping GTTC reduced infectiousness. In other words, starting GTTC might have produced the second peak, whereas ceasing GTTC produced the third peak, which suggests that the GTTC effects depend on climate conditions. If so, then climate conditions can be inferred as the main factor driving the outbreak. The GTTC effects might be supplemental. In fact, even through the end of November, GTTC significantly decreased the effective reproduction number controlling climate conditions and mobility [6]. If one extends the considered period to include the end of December, then the time at which the third peak occurred and at which GTTC ceased, then the suppressive effect of GTTC on infectiousness might be weaker or might disappear entirely.

Alternatively, GTTC itself might have had no effect on infectiousness. In fact, news\ media reports about starting or ceasing GTTC might have stimulated a rise in risk perception among the general population and might have induced more precautionary behaviors such as more scrupulous mask wearing, maintaining social distance, and cancellation of group dining. In other words, persons at leisure venues might feel higher risk from tourists; tourists might also feel higher risk of infecting others by starting GTTC. Ceasing GTTC might then induce feelings of even higher risk among the general population.

Actually, some countermeasures in addition to those considered in the present study were used, such as quarantine, isolation, PCR testing, treatment, and vaccine and drug administration. However, to examine their specific effects on infectiousness at a community level, the three considered countermeasures might be the most important policies. Moreover, the periods of the three countermeasures were not overlapped. Therefore, we can isolate the effects of the respective countermeasures more easily.

Among efforts undertaken in Japan, GTTC was one of the “Go To Campaigns (GTC).” Measures supporting GTC include “Go To Eat”, which subsidized customer bills at restaurants from the end of September through November 24 and “Go To Shopping Malls”, which subsidized shopping malls to encourage events, product development, and PR activities from October 19, 2020 through January 11, 2021, other than GTTC. However, because these were similar subsidy policies and because their periods overlapped, we cannot distinguish each branch of GTC separately. Of those, GTTC was the first and longest campaign among GTC. It was inferred as the main cause of the third wave [1]. We examined the effects of GTTC as a representative of GTC, on infectiousness. In this sense, the effects of GTTC might be the effect of GTC.

The present study has some limitations. First, R(*t*) represents infectiousness, but not in the number of patients or cases of mortality. One must be reminded that temperature and mobility are associated with infectiousness, but the result does not reflect association with the number of patients. To assess such an association, a formal mathematical model incorporating temperature and mobility must be developed. Producing that model is anticipated as a challenge for future research.

Secondly, readers must be reminded that our obtained results do not indicate some causality when interpreting the obtained results. We proved that some association exists among games with audience and lower infectiousness. That finding does not necessarily mean that games with an audience lead to lower infectiousness. It might imply that lower infectiousness, for instance, an effective reproduction number being smaller than one, induces emergency status or GTTC.

## Conclusion

Results obtained from this study demonstrated that games with an audience might not raise infectiousness. Moreover, GTTC was found to have not raised infectiousness. These results might advocate the value of holding the Olympic and Paralympic Games with audiences. Even so, the audience size must be limited to be half of the capacity of the stadium or 5,000 persons.

The present study is based on the authors’ opinions: it does not reflect any stance or policy of their professionally affiliated bodies.

## Data Availability

Japan Ministry of Health, Labour and Welfare. Press Releases

https://www.mhlw.go.jp/stf/seisakunitsuite/bunya/kenkou_iryou/kenkou/

## Acknowledgments

We acknowledge the great efforts of all staff at public health centers, medical institutions, and other facilities who are fighting the spread and destruction associated with COVID-19.

## Ethical considerations

All information used for this study was published data on the web were used. There is therefore no ethical issue related to this study.

## Competing Interest

No author has any conflict of interest, financial or otherwise, to declare in relation to this study.

## Notes

### Competing Interest Statement

The authors have declared no competing interest.

### Funding Statement

The author(s) received no specific funding for this work.

### Author Declarations

All information used for this study has been published. There is therefore no ethical issue related to this study.

### Summary of Updates

I focused on the effect of ban event so as to consider the risk of Olympic games in the same framework in our previous researches.

